# Factors associated with repeat childbirth among Adolescent Mothers in Soroti District, Teso sub-region, Uganda: A cross-sectional study

**DOI:** 10.1101/2022.05.05.22274737

**Authors:** Posiano Mulalu, Benon Wanume, Soita David, Dinnah Amongin, Gabriel Julius Wandawa

## Abstract

The percentage of adolescent mothers aged 15 to 19 years with a repeat childbirth in Uganda (26.1%) is higher than the global estimate (18.5%). Soroti district tops Teso (a region with highest adolescent childbearing rate nationally) in adolescent childbearing. Adolescent repeat childbearing (ARC) is associated with poor health outcomes, increased risk of stillbirth, maternal and child mortality, thus a public health concern. The factors associated with, and the burden, of ARC remains unknown in Soroti district. Consequently, interventions which combat ARC in Soroti district have not been informed by empirical data. This study determined the proportion of adolescent mothers with, and factors associated with, repeat childbirth among adolescent mothers in Soroti district.

We conducted a cross-sectional study involving mixed methods of data collection. Interviewer-administered structured interviews were conducted amongst 422 adolescent mothers.

Demographic and socio-economic data of respondents, data regarding respondents’ family and peer related factors was collected. Chi-square was the test statistics used. Multivariate analysis was by logistic regression.

Of the 422 respondents, 31.28% (132) were married. Proportion of respondents with repeat childbirth was at 30.81% (95%CI: 26.57%-35.39%). Risk factors of ARC were (a) being married, AOR 5.74 (95%CI: 3.08-10.68), (b) incorrect knowledge of rhythm method, AOR 2.15 (95%CI: 1.21-3.82), (c) Age at first birth, AOR 0.48 (95%CI: 0.36-0.63). Protective factors of ARC included (a) not drinking alcohol, AOR 0.41(95%CI: 0.21-0.77), not being raped, AOR 0.19(95%CI: 0.06-0.55), having first childbirth from health facility, AOR 0.38 (95%CI: 0.18-0.78) and father of first baby without multiple sexual partners, AOR 0.40(95%CI: 0.22-0.72).

In conclusion, sexual partner characteristics were associated with ARC suggesting male involvement (for example in family planning) in prevention of ARC. In addition, strengthening the implementation of anti-teen marriage programs and alcohol consumption policies, and instating measures to delay age at first delivery among adolescent mothers are required. Further research is needed to validate these findings.

## Introduction

Adolescent repeat childbearing (ARC) remains a global public health challenge, resulting into poor health, social and economic outcomes [1–4]. World Health Organization (WHO) defines an adolescent as a person aged 10 to 19 years of age[5]. Adolescent repeat childbearing (ARC) is defined by WHO as a second (or more) pregnancy ending in a live birth before age of 20 years [6, 7].

In 2018, it was estimated that, 44 live childbirths occurred among every 1000 girls aged 15 to 19 years globally [8]. The percentage of adolescent mothers aged 15 to 19 years of age with a repeat childbirth globally was estimated at 18.5% in 2017[9]. Worldwide, high rates of adolescent childbearing have been cited in developing countries. Approximately sixteen million girls aged 15 to 19 years give birth annually in developing countries [1, 10]. Since sub-Saharan Africa has the lowest contraceptive use globally, its adolescent childbearing rate (ACR) may rise, unless addressed [11].

There exist variations in the percentage of adolescent mothers aged 15 to 19 years of age with a repeat childbirth within the African countries, ranging from 3.7% in Rwanda to 26.1% in Uganda[9].

Nationally, 25% of Uganda’s adolescent girls have begun child bearing[12]. Higher ACR exist in rural areas of Uganda; 27% compared to the 19% in urban areas[12]. Uneducated Ugandan adolescent girls have a higher ACR estimated at 35%, compared to their counterparts who have attained Post-Primary Education, whose estimate is at 11%[12]. Adolescent childbearing is more common among adolescents from low wealth quintile, estimated at 33.5%, than adolescents from highest wealth quintile, whose childbearing rate is estimated at 15.1% [12]. In Uganda, Teso region has the highest ACR, estimated at 31.4% [12].

In Uganda, fluctuations exist in the estimation of adolescent repeat childbearing rate (ARCR). Some studies have reported 55.6% while others 26.1%[9, 13].

Childbearing among adolescent girls is associated with poor health outcomes among the adolescent girls and their children [14–16]. However, repeat adolescent childbirths are associated with more health risks for the mother and her child including increased risk of eclampsia, puerperal sepsis, systemic infections, low birth weight, preterm delivery and severe neonatal complications [6, 14, 17, 18]. Pregnancy and Adolescent childbirth complications are the leading cause of death among adolescent girls aged 15 to 19 years in developing countries [10]. Infants born as repeat birth to an adolescent girl are usually too small or too soon, increased child mortality and maternal mortality [19–23].

When an adolescent girl get more than one live birth, her potential to finish her education or even to get a job is limited [19]. Often, as adolescent girls move into new sexual relationship with the father of their second baby, there is a high likelihood of HIV/AIDs acquisition. Evidence shows that repeat pregnancies among adolescent girls are associated with high HIV prevalence compared to first time adolescent pregnancies [24]. Also, the first babies of the adolescent girls are often deprived of parental care, as the adolescent girls alter sexual partners. This negatively impacts on the child development[25]. Adolescents mothers with more than one gestation have lower self-esteem, greater susceptibility to unplanned pregnancy and increased use of drugs[26].

High ARCR coupled with the broad based nature of Uganda’s population pyramid is likely to have a negative impact on the Social, Health, and Economic indices of Uganda [27]. In the year 2014, 47.9% of Uganda’s population was aged zero to fourteen years (Uganda Bureau of Statistics, 2014b). This means that the above age group has either transformed into adolescents or is sooner transforming into adolescents. Adolescence is a very challenging stage, more especially in Uganda where half of her girls aged 15 to 19 years have ever had sex [29]; with the median age for sexual activity being 16.7 years [30]. Sometimes adolescent girls have reported men ten years older than them, as their sexual partners at the first sexual encounter [30]. Yet at-times such men have multiple sexual patterns [29]. This increases the risk of acquiring sexually transmitted infections (STI), pregnancy and its other associated outcomes among adolescent girls. The median birth Interval of twenty two months in Uganda [31] implies a high likelihood of Ugandan adolescent girls to have their repeat childbirth while still in adolescence. As a result, Uganda’s healthcare system is likely to be overwhelmed with health issues linked to ARC. Therefore, there is urgent need to prevent ARC so as to achieve the Sustainable Development Goal (SDG) number three (ensure healthy lives and promote well-being for all at all ages)[32].

Adolescent repeat childbearing (ARC) has attracted global and national attention. The Uganda government in collaboration with several non-governmental Organizations has intervened in various ways, including; development of National Adolescent policy, expansion and strengthening of youth friendly sexuality related services, enacting of laws and policies relating to adolescent sexuality, such as the defilement act, anti-teen marriage act, among others [2, 33, 34].

Despite the above interventions, a slight decline ARCR in Uganda has been seen; 58.9% in 1988/89 to 55.6% in 2016 [13]. This could be attributed to challenges faced by policy and program implementation [35]. Such challenges may include; cultural influence in the local areas [35]. Therefore, precisely, to scale down adolescent repeat childbearing rate in Soroti district, evidence based interventions (interventions informed by the burden of, and factors associated with, ARC in the district) are required.

## Methods and Materials

### Study area/study setting

The study was conducted in Soroti district. Soroti district has the highest percentage of adolescent mothers who have begun childbearing within Teso sub region “Table 1”.

**Table 1.**
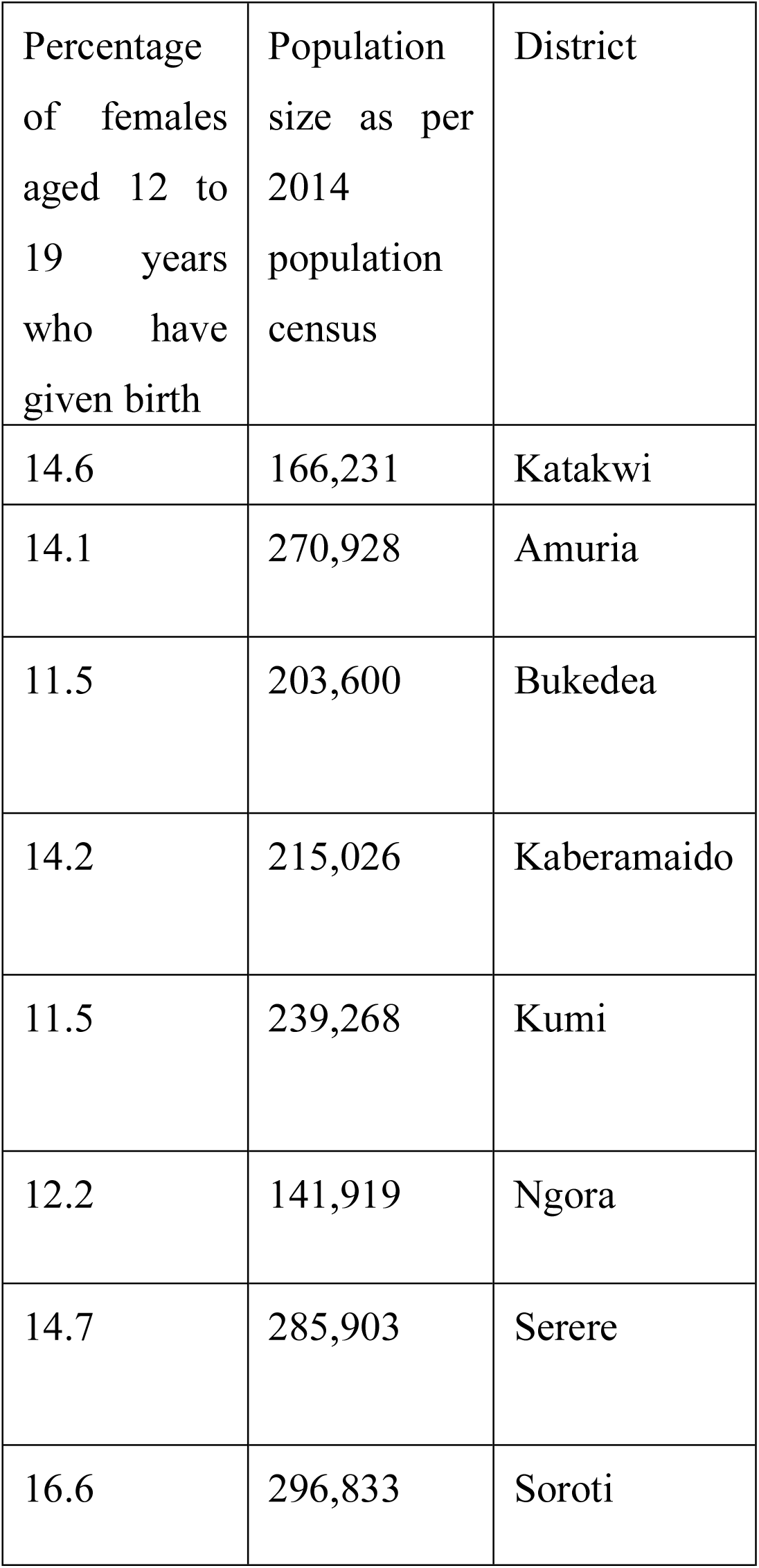
showing previous child bearing rates among adolescent females in various districts of Teso region.

The socio-cultural context in Soroti district perpetuates a high population size of people of various socio-demographics (population size of 296,833 people in 2014, of which, 13.7% are adolescent girls[28, 36]). Soroti district has an area of 2662Km. Kaberamaido, Amuria, Katakwi and Serere are its neighboring districts in the west, east, north and south respectively “Fig 1”[28]. Soroti district has three constituencies, 10 sub-counties, and 331 parishes. The major ethnic groups in Soroti are the Itesots and Kumams [28].

**Fig 1.**
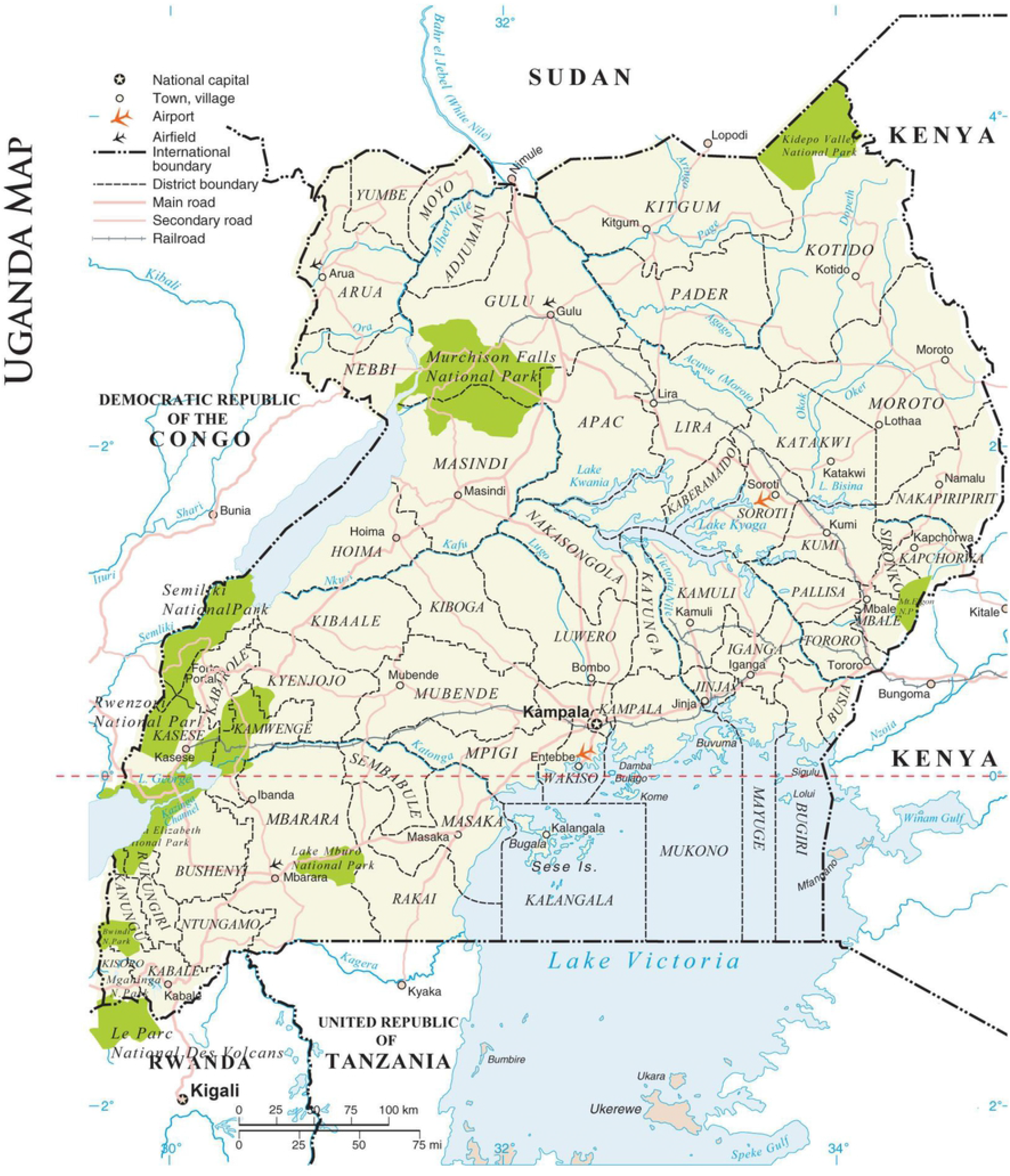
Map of Uganda showing study area.

### Study design

The study involved a cross-sectional study was used to capture data on a set of factors related to adolescent repeat childbirth.

Numerical data was collected on predictive factors of repeat childbirth among 422 adolescent mothers aged 15 to 19 years in Soroti district using an interviewer administered questionnaire.

### Study Population

Study population were adolescent mothers aged 15 to 19 years, with a repeat childbirth in Soroti district.

### Sample size

Kish Leslie’s proportion formula was used to calculate the sample size.

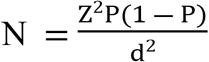 N= Sample size, where Z = 1.96; the statistical standard normal variate at 5% significance level, d = 0.05; the precision level, p=rate of repeat childbearing among adolescent mothers.

Since the repeat childbearing rate among adolescent mothers in Soroti district was unknown by then, the rate of repeat childbearing among adolescent mothers for Uganda was used. Therefore, the value of p was 55.6% % [12].

Using the above values, the computed sample size was 380 participants.

Taking into consideration failure to complete questionnaire rate of 10%, the sample size was 422 participants.

### Sampling procedure

Two out of three counties (Dakebala and Soroti County) were randomly selected to participate in the study. This was done by writing the names of all counties on separate pieces of papers, folding and drawing them in a single container then only two counties were picked at the same time. At county level, all sub-counties were selected to participate in the study since data regarding adolescent child bearing rate at sub-county had not been documented. At sub-county level all parishes and villages were included in the study. At village level, the list of households (HHs) was developed by researcher with the help of Local council one (LCI) chairperson. Then, the first household to participate was selected by simple random sampling technique from the many HHs within the village. This was done to ensure that each household had an equal chance of being selected. The study households which are preceded by the first household were selected using a systematic sampling technique. If a selected household did not have an adolescent mother; the immediate neighboring household was selected. This was continued until the required number of participants was obtained. In case a given household had more than one adolescent mother, only one adolescent mother was selected using simple random technique; where their names were written on separate pieces of papers, and then folded, and mixed up and one selected at random. The above procedures were followed until the required number of participants was obtained in each county.

#### Inclusion Criterion

Adolescent mother aged 15 to 19 years within the selected household (if a household had only one adolescent mother) who consented to participate in the study

Randomly selected adolescent mother aged 15 to 19 years within the selected household (if a household had more than one adolescent mother) who consented to participate in the study

#### Exclusion criterion

Adolescent mother aged 15 to 19 years within the selected household (if a household had only one adolescent mother) and she failed to consent to participate in the study was excluded.

Apart from the selected adolescent mother, other adolescent mothers aged 15 to 19 years within the selected household (if a household had more than one adolescent mother) were excluded.

Adolescent mothers who lost their babies were also excluded. Visiting adolescent mothers were excluded.

Adolescent mothers who were reported to be mentally impaired also excluded.

### Data collection tool

An interviewer-administered questionnaire was administered to selected adolescent mother who consented to participate.

### Data Collection methods

Research assistants administered questionnaires to selected participants at the selected households. The research assistants were trained prior to data collection on different aspects of the questionnaire.

Data collection for the quantitative component lasted for three weeks.

### Study validity

#### Internal validity

Consistent questionnaires were used. The questionnaires were pretested. Consistent sampling procedures were also used throughout the study. Potential confounders, effect modifiers and bias were controlled for at study design, data collection and at analysis level through using multiple logistic regressions.

There was translation of study into local languages (for participants who did not understand English) and re – translation back to English, was done by research assistants.

#### External validity

External validity was achieved through calculation of appropriate representative sample size, use of explicit study design and appropriate statistical analytical tools.

### Data Management

#### During and after data collection

The research assistant administered the questionnaires to the participant. Before the research assistant would administer another questionnaire to the next participant, the research assistant checked if she/he had fully administered all the questions in the questionnaire to the participant. If there were questions that had not been administered, the research assistant would administer those questions to the participant before the participant left.

The Principal Investigator equally checked the Questionnaire for completeness before acceptance from the Research Assistant.

Data from completed Questionnaires was entered into Excel sheet, saved as a cvs file and then imported to Stata Version 14 for further data management and analysis.

Before data analysis, there was value labeling, variable renaming and data manipulation where possible.

Data was explored to check for consistence and outliers. Influential outlying observations were dropped.

Multiple Soft copies of the data was made and stored on different hard discs and each was kept under tight privacy and confidentiality using password.

Hard copies of the Questionnaires were safely kept in locked cardboards.

#### Data Management during data processing

Data was cleaned for errors and omissions by comparing what was in the computer database with what was on the hard copies, then was exported to STAT version 14 (V14), sorted, categorized and all the required variable transformations done. Caution was taken throughout the analysis. Analyzed data was kept in the sub-folder and each was encrypted for security. Final results were merged into a master file. All files were encrypted for security purposes.

#### Participant Privacy

Codes were given to participants’ questionnaires to serve as unique identifiers on the Questionnaires and NOT any personally identified information.

#### Data Quality

At data collection level and sampling level, Research assistants were trained in data collection including how to seek for consent from participants. The research assistants were supervised throughout the entire data collection period. During the data collection periods, Questionnaires were reviewed for completeness, errors and omissions. The Questionnaire was pretested to assess its reliability, validity, reliability and appropriateness prior to start of the data collection exercise.

### Data Analysis Plan

#### Proportion of adolescent mothers with repeat childbirth

This was computed as a proportion. The numerator was; total number of adolescent mothers with repeat childbirth(s). The denominator was; total number of adolescent mothers. The confidence interval was set at 95% level of confidence.

#### Factors associated with repeat childbirths among adolescent girls

The factors associated with repeat childbirths among adolescent mothers were analyzed at Univariate, Bivariate and multiple levels. Six measures of income used in the 2014 Uganda Population and housing census were used to measure income[28]. These were possession of a radio, Television, a phone, sofa set, bed, and a chair. A score of two out of six was considered low income, a score of three or four out of the six was categorized medium income and a score of five and above out of the six was high income. At bivariate level, a chi-square was used to test for significant association between repeat childbirth and each of the independent variables. The level of confidence was set at 95%. In cases, where at-least a cell had a count of 5 or less, Fisher’s exact test was used to test for significant association between the outcome and a given independent variable. Influential outliers among the observations were dropped (11 observations). An outlier was considered influential when the magnitude of its standard residuals (|stdres| >= 2) was equal to or greater than two and the magnitude of its influential residues (|infres|>=7) was equal to or greater than seven. These observations were dropped to avoid their effect on the model estimation. The multi-collinearity was tested for among the independent variables. Factors that showed no significant association with repeat childbirth but were significantly associated with repeat childbirth from other related studies (biological plausibility) and had p<0.25 from the current study, were also used during the model building. The best model fit was then selected “Fig 2”.

**Fig 2.**
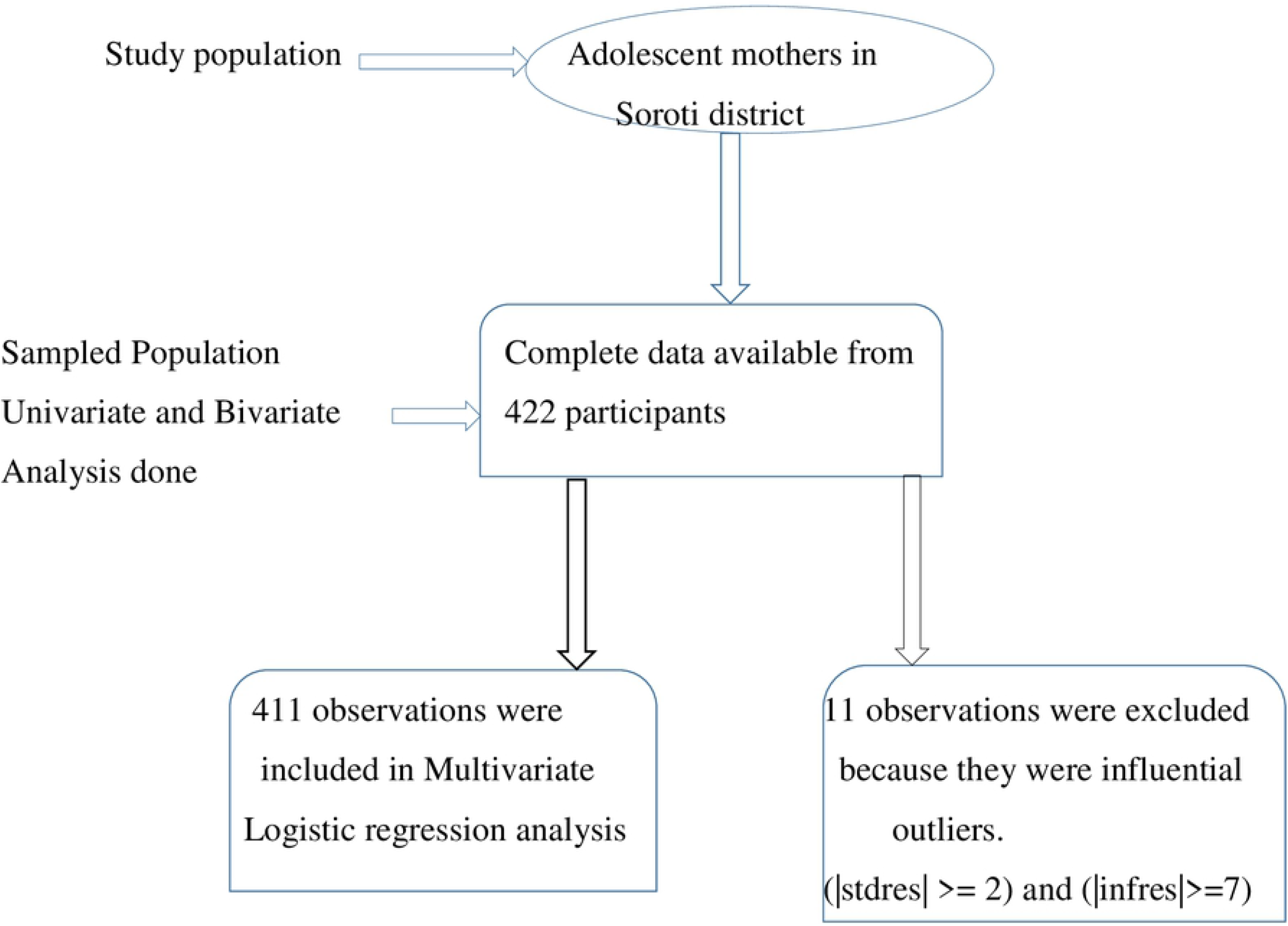
Data Analysis flow chart.

A contingency table stating the frequencies of each option per variable was made. The frequencies of each outcome of a given variable were represented graphically. The descriptive statistics of each variable was analyzed and tabulated. The statistically significant factors were chosen and reported. The order of entry of the variables into the table was by default; based on the way variables were entered the STAT v14 software.

### Data Presentation

Study findings were presented inform of tables and graphs.

## Ethical consideration

Ethical clearance was sought from Mbale Regional Referral Hospital Research and Ethics Committee. Permission was thought from the DHO of Soroti district. Clearance was obtained from Busitema University, and higher degrees committee of Busitema University.

Photocopies of the approval letter from the Research and Ethics Committee and introductory letter from Dean faculty of Health sciences, Busitema University was made and each separately counter-signed by the District Health Officer Soroti district. These letters were further photocopied, and the photocopies used as introductory letters to the different leaders at various levels.

A clear and detailed description of the study protocol was given to the different stakeholders including, the leaders and study participants. An informed consent was obtained from the study participants since some who were below 18 years were emancipated minors since they had given birth.

Equally, the local council chairpersons of the villages were approached by the researcher.

They were be explained to, the research details. Formal permission from the local council chairpersons was sought, through obtaining oral consent.

Data was collected from participants who consented to participate. High degree of professionalism, confidentiality and privacy was exhibited during the data collection exercise.

Participants’ names were not collected during the data collection processes, data analyses and research publication. Rather, codes were used to ensure participant confidentiality and anonymity.

Furthermore, the completed participant consent forms were kept under locked cupboards and only accessible to the study team or authorized person(s) in case need arises.

## Dissemination of results

Results of the study have been availed to Busitema University, the District Health Office of Soroti, and Mbale RRH REC and orally presented to area gatekeepers/leaders from which data was collected.

### Study Limitations

Recall and Information bias were the likely limitations of the study since participants were asked about past events in their life.

External validity is a limitation to the findings of the qualitative data. Findings can only be applied to Soroti district, Teso sub-region.

### Psycho-social Support

No participant had her deep-seated emotions awakened.

### Safety of study team and Participants against COVID19

There was strict adherence to the ministry of health standard operating procedures against COVID 19. Social distancing of at-least two meters and use of hand sanitizers and masks were strictly worn by the study team.

## RESULTS, INTERPRETATION AND PRESENTATION

### Proportion of adolescent mothers with a repeat child birth

Out of the 422 respondents, 30.81% (n=130; 95% CI: 26.57% - 35.39%) reported to have a repeat childbirth “Table 2”.

**Table 2.** Proportion of adolescent mothers with repeat childbirth.

| Variable | Frequency | Proportion | Standard Error | [95% Confidence Interval] |
| --- | --- | --- | --- | --- |
| Single childbirth | 292 | 0.6919431 | 0.0225014 | [0.6460508 - 0.734332] |
| Repeat child birth | 130 | 0.3080569 | 0.0225014 | [0.265668 - 0.3539492] |

### Demographic, socio-economic and sexual/reproductive characteristics of Participants

The study was carried out in two constituencies, representing 66.67% of the district’s constituencies. The mean age of the 422 respondents was 18.27, with a standard deviation of 0.97, and their median age was 18 years. Of the total number of participants, 78.44% were aged 18 to 19 years, 52.13% were Anglicans, 94.79% were from rural areas, and 68.72% were unmarried. 69.19 % were Itesots, 69.19% ended in primary and 72.99% were not engaged in any income generating activity.

Above eighty percent (84.12%) of the respondents were staying with their parents by the time they had their first childbirth, 83.89 % were of the same tribe as the fathers of their first baby, 76.78% were at school at the time of their first pregnancy, 84.36% delivered their first baby from health facility, 89.81% were HIV negative, 67.30% had correct knowledge of calendar family planning method, 71.33% did not use condoms during their last sexual encounter, 47.63% could not insist on condom use if their sexual partner refused, 89.34% believed that other contraceptive methods including pills could prevent pregnancy, 18.72% drink alcohol and 5.45% were raped 40.05% were physically assaulted.

#### Family and peer related characteristics

Above seventy eighty percent (78.44%) of the participant had both parents alive, 78.67% had their fathers as the family head, 31.04% of the respondents reported their mothers to have had a first childbirth when they were 19 years of age or less, and 71.09% came from low income families.

Above sixty three percent (63.74%) of the participants had peers with a repeat childbirth. 37.44% of the participants were encouraged by their peers to have a repeat childbirth “Table 6”.

#### Sexual partner (father of respondent’s first baby) related characteristics

Of the total number of participants, 50.71% had the fathers of their first babies aged 20 to 24 years, 54.50% of the respondents’ sexual partners were Anglicans, 32.23% of the sexual partners of the respondents were married to the respondents, 33.41% of the participants’ sexual partners were married to other women, 34.83% of the sexual partners of the respondents had other children, 62.32% of the participants’ sexual partners ended in primary, 66.35% of the participants’ sexual partners were low income earners, and 73.22% of the participants had the fathers of their first babies HIV negative.

### Association between participant’s socio-demographics and repeat childbirth

The association between the various factors and repeat childbirth was determined using a 2×2 table and the Pearson Chi-square or Fisher’s exact tests (if a variable had at least a cell with a value of at-most 5). Furthermore, respondents’ age was also dichotomized using the median age. A significant association arose between respondent’s age group (X^2^=9.52, p=0.002), marital status (X^2^=28.17, p=0.000), Tribe (X^2^=12.46, p=0.002), occupation (X^2^=4.45, p=0.035) and mother’s age at first birth (p=0.000) with repeat childbirth “Table 3”.

**Table 3:**
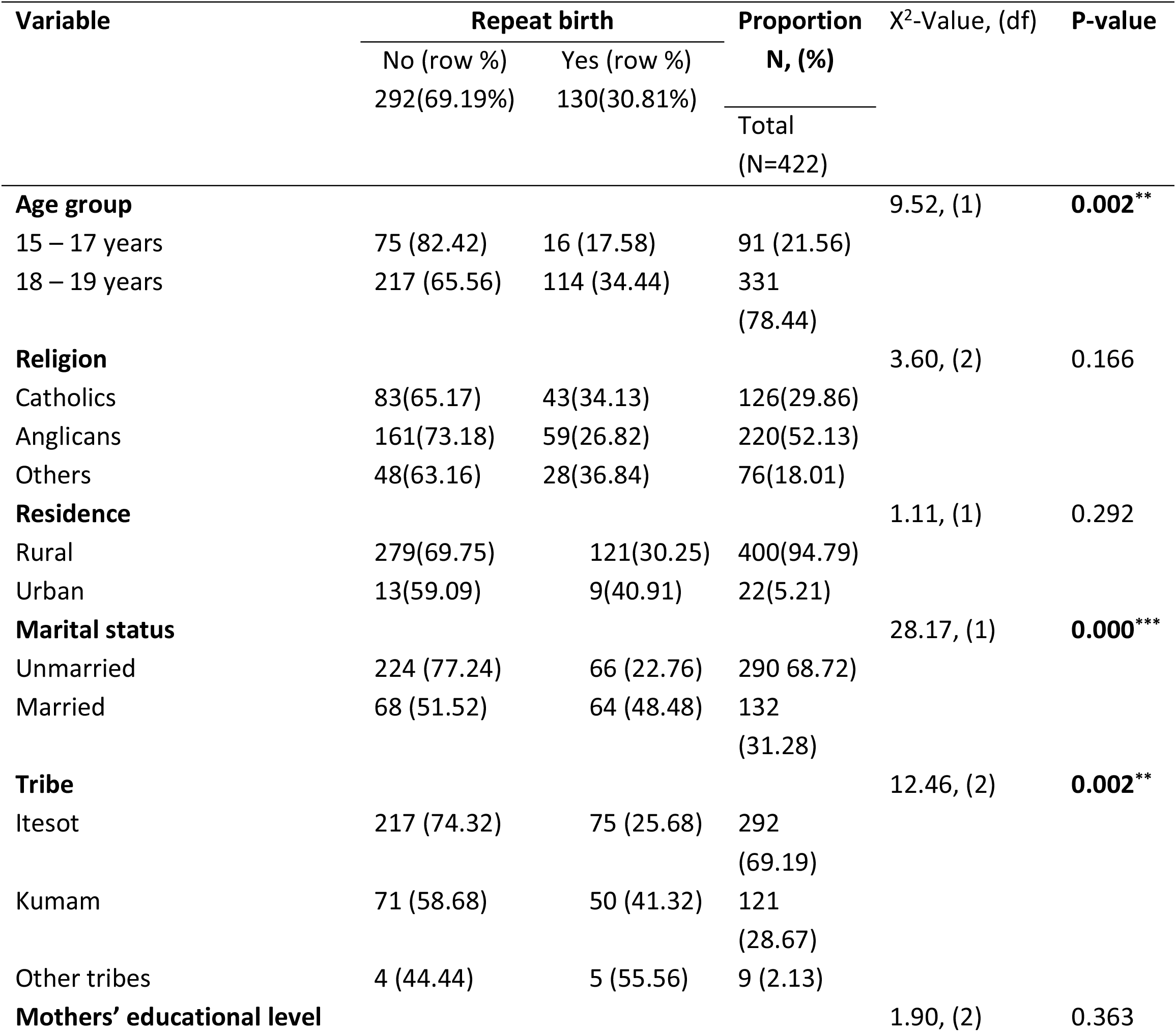
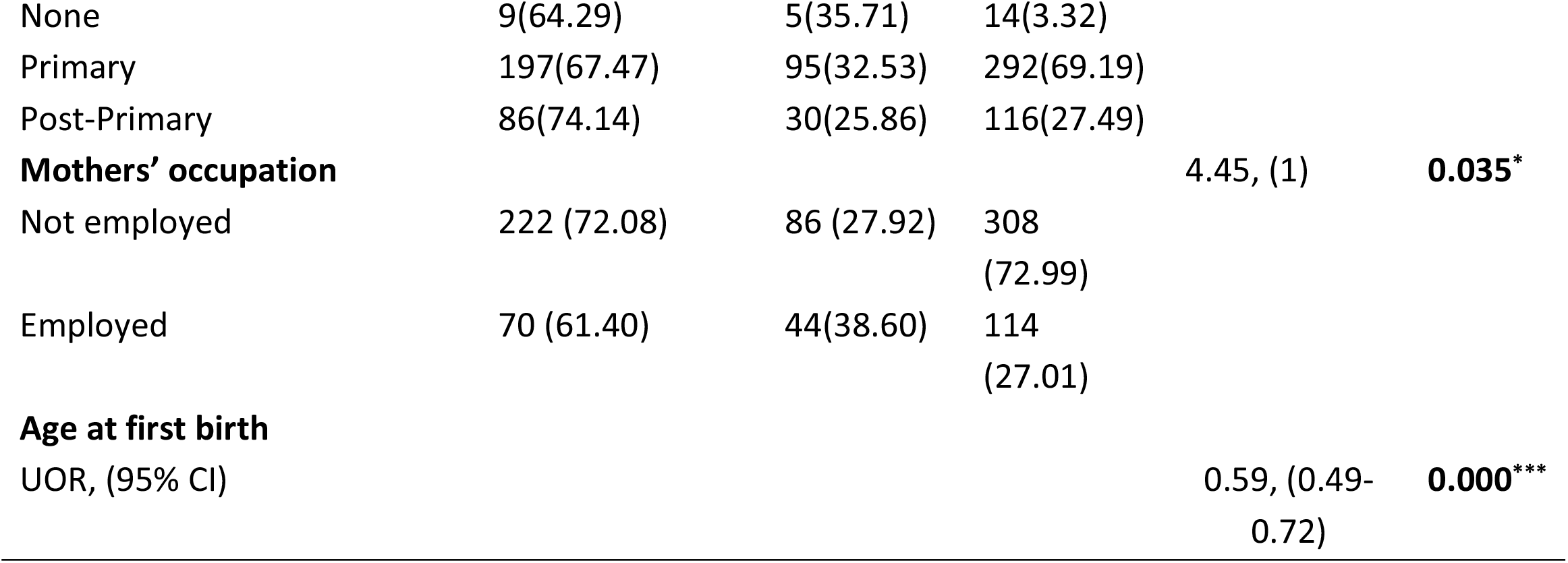
Association between mother’s socio-demographics and repeat childbirth.

| Variable | Repeat birth |  | Proportion<br>N, (%) | X<sup>2</sup>-Value, (df) | P-value |
| --- | --- | --- | --- | --- | --- |
|  | No (row %) | Yes (row %) |  |  |  |
|  | 292(69.19%) | 130(30.81%) | Total<br>(N=422) |  |  |
| **Age group** |  |  |  | 9.52, (1) | **0.002\*\*** |
| 15 – 17 years | 75 (82.42) | 16 (17.58) | 91 (21.56) |  |  |
| 18 – 19 years | 217 (65.56) | 114 (34.44) | 331<br>(78.44) |  |  |
| **Religion** |  |  |  | 3.60, (2) | 0.166 |
| Catholics | 83(65.17) | 43(34.13) | 126(29.86) |  |  |
| Anglicans | 161(73.18) | 59(26.82) | 220(52.13) |  |  |
| Others | 48(63.16) | 28(36.84) | 76(18.01) |  |  |
| **Residence** |  |  |  | 1.11, (1) | 0.292 |
| Rural | 279(69.75) | 121(30.25) | 400(94.79) |  |  |
| Urban | 13(59.09) | 9(40.91) | 22(5.21) |  |  |
| **Marital status** |  |  |  | 28.17, (1) | **0.000\*\*\*** |
| Unmarried | 224 (77.24) | 66 (22.76) | 290 68.72) |  |  |
| Married | 68 (51.52) | 64 (48.48) | 132<br>(31.28) |  |  |
| **Tribe** |  |  |  | 12.46, (2) | **0.002\*\*** |
| Itesot | 217 (74.32) | 75 (25.68) | 292<br>(69.19) |  |  |
| Kumam | 71 (58.68) | 50 (41.32) | 121<br>(28.67) |  |  |
| Other tribes | 4 (44.44) | 5 (55.56) | 9 (2.13) |  |  |
| **Mothers' educational level** |  |  |  | 1.90, (2) | 0.363 |
| None | 9(64.29) | 5(35.71) | 14(3.32) |  |  |
| Primary | 197(67.47) | 95(32.53) | 292(69.19) |  |  |
| Post-Primary | 86(74.14) | 30(25.86) | 116(27.49) |  |  |
| **Mothers' occupation** |  |  |  | 4.45, (1) | **0.035\*** |
| Not employed | 222 (72.08) | 86 (27.92) | 308<br>(72.99) |  |  |
| Employed | 70 (61.40) | 44(38.60) | 114<br>(27.01) |  |  |
| **Age at first birth** |  |  |  |  |  |
| UOR, (95% CI) |  |  |  | 0.59, (0.49-0.72) | **0.000\*\*\*** |

### Association between other factors of participant and repeat childbirth

A significant association was observed between place of delivery of first baby (X^2^=9.59, p=0.002), knowledge of family planning (calendar method) (X^2^=13.73, p=0.000), alcohol consumption (X^2^=13.64, p=0.000), rape (X^2^=5.21, p=0.022) and physically assault (X^2^= 7.75, p=0.005) with repeat childbirth. Of the respondents with repeat child births, half were physically assaulted, 23.85% delivered from home, 45.38% did not have correct knowledge about calendar family planning method, 29.23% drink alcohol, and 9.23% had been raped “Table 4”.

**Table 4:** Association between other factors of participant and repeat childbirth.

| Variable | Repeat child birth |  | Proportion<br>N, (%)<br>Total(N=422) | X<sup>2</sup>-<br>Value,<br>(df) | P-value |
| --- | --- | --- | --- | --- | --- |
|  | No (row %)<br>292(69.19%) | Yes (row %)<br>130(30.81%) |  |  |  |
| **At time of first delivery, participant was staying with** |  |  |  | 1.56(2) | 0.458 |
| Parents | 249(70.14) | 106(29.86) | 355(84.12) |  |  |
| Alone | 9(56.25) | 7(43.75) | 16(3.79) |  |  |
| Relatives | 34(66.67) | 17(33.33) | 51(12.09) |  |  |
| **Participant's tribe same as first sexual partner** |  |  |  | 0.31(1) | 0.576 |
| Yes | 243(68.64) | 111(31.36) | 354(83.89) |  |  |
| No | 49(72.06) | 19(27.94) | 68(16.11) |  |  |
| **Participant was schooling at time of her first pregnancy** |  |  |  | 0.20(1) | 0.651 |
| Yes | 226(69.75) | 98(30.25) | 324 (76.78) |  |  |
| No | 66(67.35) | 32(32.65) | 98(23.22) |  |  |
| **Site of delivery of first baby** |  |  |  | 9.56(1) | **0.002\*\*** |
| Home | 35(53.03) | 31(46.97) | 66 (15.64) |  |  |
| Health facility | 257 (72.19) | 99 (27.81) | 356 (84.36) |  |  |
| **HIV status of participant** |  |  |  | 0.07(2) | 0.958 |
| Negative | 263(69.39) | 116(30.61) | 379(89.81) |  |  |
| Positive | 6(66.67) | 3(33.33) | 9(2.13) |  |  |
| Unknown | 23(67.65) | 11(32.35) | 34(8.06) |  |  |
| **Knowledge of Calendar Family planning method** |  |  |  | 13.73(1) | **0.000\*\*\*** |
| Knows | 213(75.00) | 71(25.00) | 284(67.30) |  |  |
| Don't know | 79(57.25) | 59(42.75) | 138(32.70) |  |  |
| **Participant used Condom during the last sexual encounter** |  |  |  | 0.75(1) | 0.385 |
| Yes | 80(66.12) | 41(33.88) | 121(28.67) |  |  |
| No | 212(70.43) | 89(29.57) | 301(71.33) |  |  |
| **Can insist on Condom use** |  |  |  | 0.00(1) | 0.986 |
| Yes | 153(69.23) | 68(30.77) | 221(52.37) |  |  |
| No | 139(69.15) | 62(30.85) | 201(47.63) |  |  |
| **Effectiveness of Family planning methods** |  |  |  | 0.09(1) | 0.768 |
| Yes | 260(68.97) | 117(31.03) | 377(89.34) |  |  |
| No | 32(71.11) | 13(28.89) | 45(10.66) |  |  |
| **Drinks alcohol** |  |  |  | 13.64(1) | **0.000\*\*\*** |
| Yes | 41(51.90) | 38(48.10) | 79(18.72) |  |  |
| No | 251(73.18) | 92(26.82) | 343(81.28) |  |  |
| **Raped** |  |  |  | 5.21(1) | **0.022\*** |
| Yes | 11(47.83) | 12(52.17) | 23(5.45) |  |  |
| No | 281(70.43) | 118(29.57) | 399(94.55) |  |  |
| **Physically assaulted** |  |  |  | 7.75(1) | **0.005\*\*** |
| Yes | 104(61.54) | 65(38.46) | 169(40.05) |  |  |
| No | 188(74.31) | 65(25.69) | 253(59.95) |  |  |

Note: \* $p < 0.05$ , \*\* $p < 0.01$ , \*\*\* $p < 0.001$ df=degrees of freedom

### Association between Participant’s family related factors and repeat childbirth

No family related factor was identified to have a significant association with repeat childbirth “Table 5”.

**Table 5:** Association between Participant’s family related factors and repeat childbirth.

| Variable | Repeat childbirth |  | Proportion<br>N, (%) | X<sup>2</sup>-<br>Value,<br>(df) | P-value |
| --- | --- | --- | --- | --- | --- |
|  | No (row %) | Yes (row %) |  |  |  |
|  | 292(69.19%) | 130(30.81%) | Total<br>(N=422) |  |  |
| **Parent's survival** |  |  |  | 2.10 (2) | 0.510 |
| Both alive | 234(70.69) | 97(2.31) | 331(78.44) |  |  |
| Only Father alive | 8(2.74) | 6(42.86) | 14(3.32) |  |  |
| Only mother alive | 43(64.18) | 24(35.82) | 67(15.88) |  |  |
| Both father and mother deceased | 7(70.00) | 3(30.00) | 10(2.37) |  |  |
| **Family head** |  |  |  | 1.45(2) | 0.542 |
| Father | 232(69.88) | 100(30.12) | 332(78.67) |  |  |
| Mother | 49(64.47) | 27(35.53) | 76(18.01) |  |  |
| Others | 11(78.57) | 3(21.43) | 14(3.32) |  |  |
| **Participant Mother's childbirth history** |  |  |  | 0.49(2) | 0.783 |
| Delivered her first baby when was <20 years | 88(67.18) | 43(32.82) | 131(31.04) |  |  |
| Delivered her first baby when was ≥20 years | 34(68.00) | 16(32.00) | 50(11.85) |  |  |
| Unknown | 170(70.54) | 71(29.46) | 241(57.11) |  |  |
| **Family income** |  |  |  | 3.48(2) | 0.149 |
| Low | 215(71.67) | 85(28.33) | 300(71.09) |  |  |
| Middle | 74(63.79) | 42(36.21) | 116(27.49) |  |  |
| High | 3(50.00) | 3(50.00) | 6(1.42) |  |  |

*df=degrees of freedom*

**Table 6:**
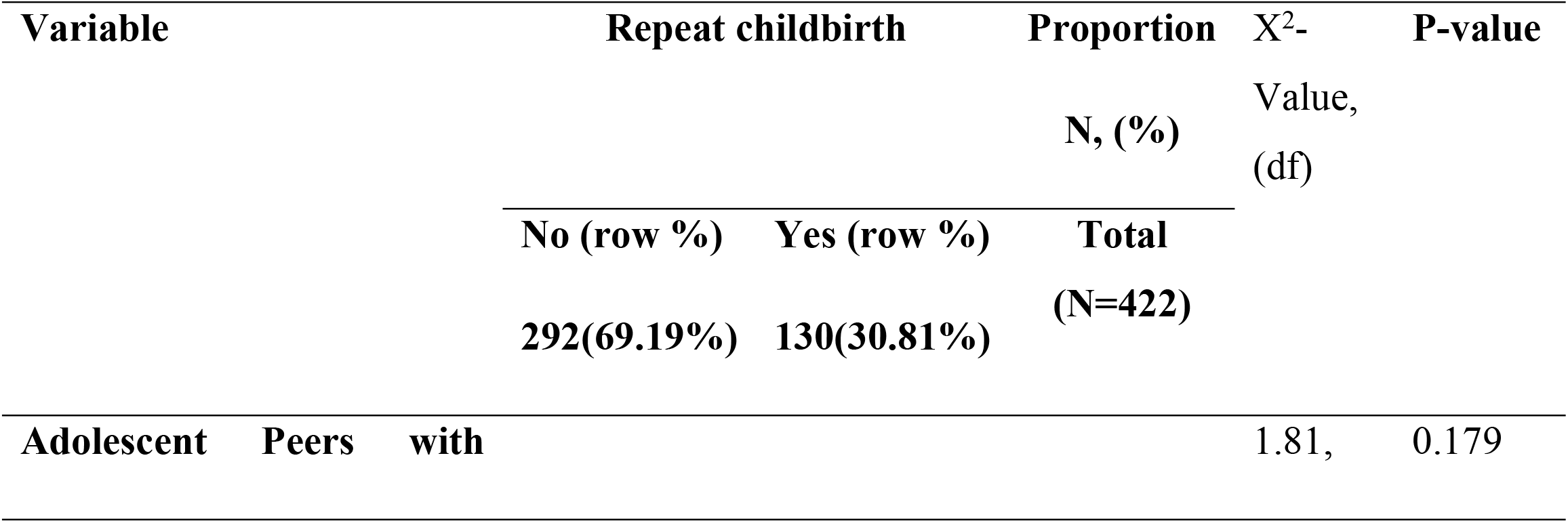
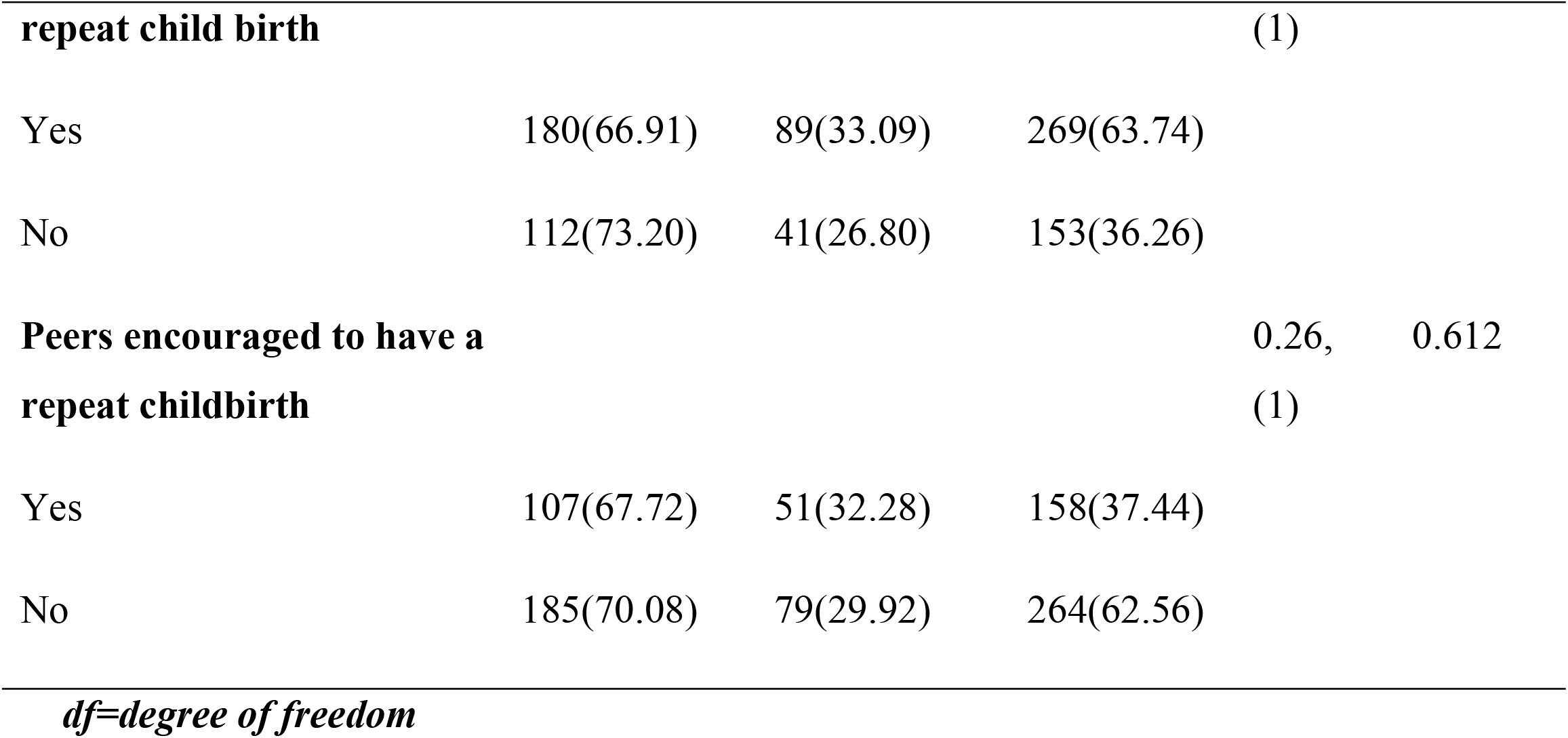
Association between participant’s peer related factors and repeat childbirth.

### Association between participant’s peer related factors and repeat childbirth

There was no significant association between peer related factors and repeat childbirth “Table 6”.

### Association between repeat childbirth and father of the respondent’s first baby (sexual partner) characteristics

A significant association was seen between the father of the respondent’s first baby being married to the respondent (X^2^=18.58, p=0.000), the father of the respondent’s first baby having multiple sexual partners (X^2^=17.22, p=0.000), the father of the respondent’s first baby having other children(X^2^=21.02, p=0.000) with repeat childbirth. Besides fathering the respondent’s first baby, most respondents reported the fathers of their first babies to have fathered children from other women. 46.92% of the respondents were married to the fathers of their first babies, and 47.69% had the fathers of their first babies with multiple sexual partners “Table 7”.

**Table 7:** Association between repeat childbirth and sexual partner’s characteristics.

| Variable | Repeat childbirth | | Proportion<br>N, (%) | $X^2$, (df) | P-value |
| --- | --- | --- | --- | --- | --- |
|  | No (row %)<br>292(69.19%) | Yes (row %)<br>130(30.81%) | Total<br>(N=422) |  |  |
| **Age group** |  |  |  | 5.38, (2) | 0.068 |
| < 20 years | 66(77.65) | 19(22.35) | 85(20.14) |  |  |
| 20-24 years | 149(69.63) | 65(30.37) | 214(50.71) |  |  |
| Above 24 years | 77(62.60) | 46(37.40) | 123(29.15) |  |  |
| **Religion** |  |  |  | 5.61, (2) | 0.135 |
| Catholics | 79(63.20) | 46(36.80) | 125(29.62) |  |  |
| Moslems | 8(80.00) | 2(20.00) | 10(2.37) |  |  |
| Anglicans | 169(73.48) | 61(26.52) | 230(54.50) |  |  |
| Other religions | 36(63.16) | 21(36.84) | 57(13.51) |  |  |
| **Married to respondent** |  |  |  | 18.58, (1) | **0.000\*\*\*** |
| Yes | 75(55.15) | 61(44.85) | 136(32.23) |  |  |
| No | 217(75.87) | 69(24.13) | 286(67.77) |  |  |
| **Multiple sexual partners** |  |  |  | 17.22, (1) | **0.000\*\*\*** |
| Yes | 79(56.03) | 62(43.97) | 141(33.41) |  |  |
| No | 213(75.80) | 68(24.20) | 281(66.59) |  |  |
| **Sexual partner has other children** |  |  |  | 21.02, (1) | **0.000\*\*\*** |
| Yes | 81 (55.10) | 66(44.90) | 147(34.83) |  |  |
| No | 211 (76.73) | 64(23.27) | 281(65.17) |  |  |
| **Education of first sexual partner** |  |  |  | 1.89, (3) | 0.538 |
| None | 6(66.67) | 3(33.33) | 9(2.13) |  |  |
| Primary | 188(71.48) | 75(28.52) | 263(62.32) |  |  |
| Secondary | 87(65.91) | 45(34.09) | 132(31.28) |  |  |
| Tertiary | 11(61.11) | 7(38.89) | 18(4.27) |  |  |
| **Income level** |  |  |  | 0.67, (2) | 0.681 |
| Low | 191(68.21) | 89(31.79) | 280(66.35) |  |  |
| Middle | 96(71.64) | 38(28.36) | 134(31.75) |  |  |
| High | 5(62.50) | 3(37.50) | 8(1.90) |  |  |
| **HIV status of sexual partner** |  |  |  | 3.19, (2) | 0.194 |
| Negative | 221(71.52) | 88(28.48) | 309(73.22) |  |  |
| Positive | 5(71.43) | 2(28.57) | 7(1.66) |  |  |
| Unknown | 66(62.26) | 40(37.50) | 106(25.12) |  |  |

*Note: \* $p < 0.05$ , \*\* $p < 0.01$ , \*\*\* $p < 0.001$ df=degree of freedom*

### Factors associated with repeat childbirth among adolescent mothers

The suitable model to explain repeat child birth was based on either; (i) factors that were significantly associated with repeat childbirth at bivariate level given that they showed no association with other independent variables or (ii) factors that showed no significant association with repeat childbirth but were significantly associated with repeat childbirth from other related studies (biological plausibility) and had p<0.25 from the current study. As a result, respondent’s education level, family income and having peers with repeat childbirth were included due to biological plausibility. Occupation of respondents and father of respondent’s first baby having other children, were dropped due to multi-collinearity with other independent variables. Influential outliers among the observations were dropped (11 observations). An outlier was considered influential when the magnitude of its standard residuals (|stdres| >= 2) was equal to or greater than two and the magnitude of its influential residues (|infres|>=7) was equal to or greater than seven. These observations were dropped to avoid their effect on the model estimation.

Respondents who had a first childbirth from a health facility are 0.38 times less likely to have a repeat childbirth as compared to respondents who delivered first baby at home (AOR 0.38 (95%CI: 0.18 – 0.78), UOR 0.42 (95%CI: 0.25-0.73). The odds of having a repeat childbirth are 2.15 times higher among participants without correct knowledge of rhythm method of family planning as compared to those with correct knowledge (AOR 2.15 (95%CI: 1.21-3.82), UOR 2.33 (95%CI:1.50-3.63). The odds of having a repeat childbirth are 5.74 times higher among married participants as compared to the unmarried (AOR 5.74 (95%CI: 3.08-10.68), UOR 3.61 (95%CI:2.30-5.65). Respondents who did not drink alcohol were 0.41 times less likely to have a repeat childbirth when compared with those who reported to drink alcohol (AOR 0.41 (95%CI:

0.21 – 0.77), UOR 0.36 (95%CI: 0.21-0.60). Respondents who reported not to have been raped were 0.19 times less likely to have a repeat childbirth when compared with those who reported to have been raped (AOR 0.19 (95%CI: 0.06 – 0.55), UOR 0.36 (95%CI: 0.16-0.85). The odds of having a repeat childbirth decreases by 52% for every one-year increase in age at first birth (AOR 0.48 (95%CI: 0.36 – 0.63), UOR 0.59 (95%CI: 0.49-0.72). Participants having the father of the first baby without multiple sexual partners are 0.40 times less likely to have a repeat childbirth as compared to participants with the father of the first baby with multiple sexual partners (AOR 0.40 (95%CI:0.22 – 0.72), UOR0.39 (95%CI: 0.25-0.60) “Table 8”.

**Table 8:**
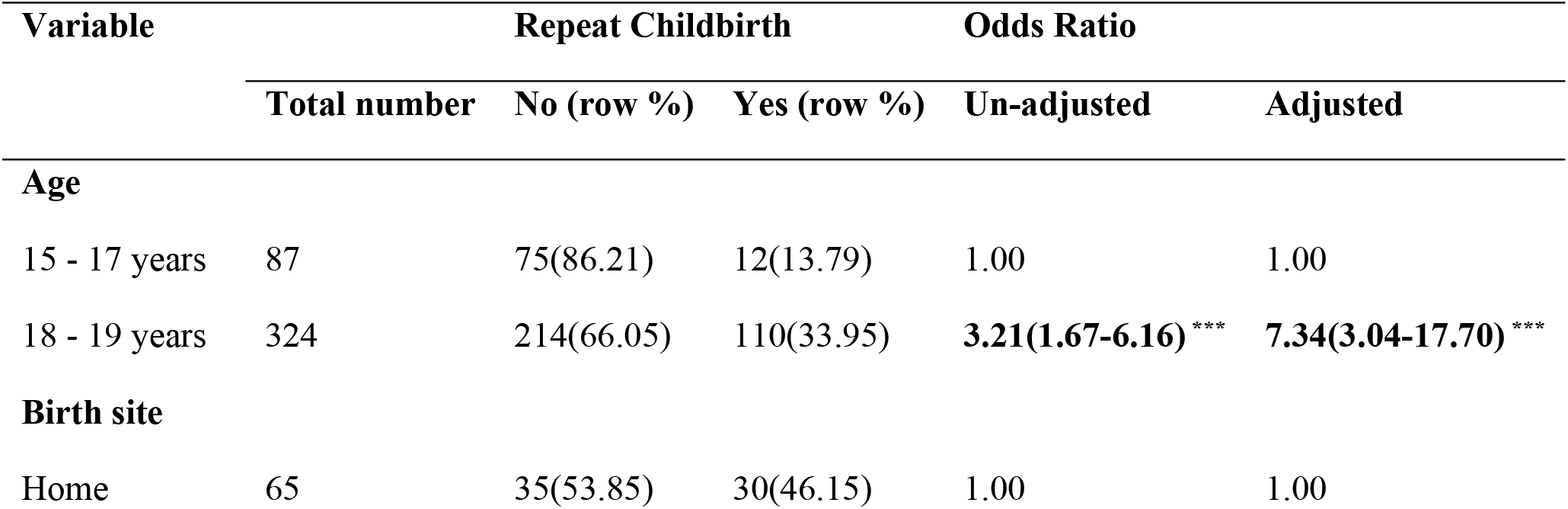
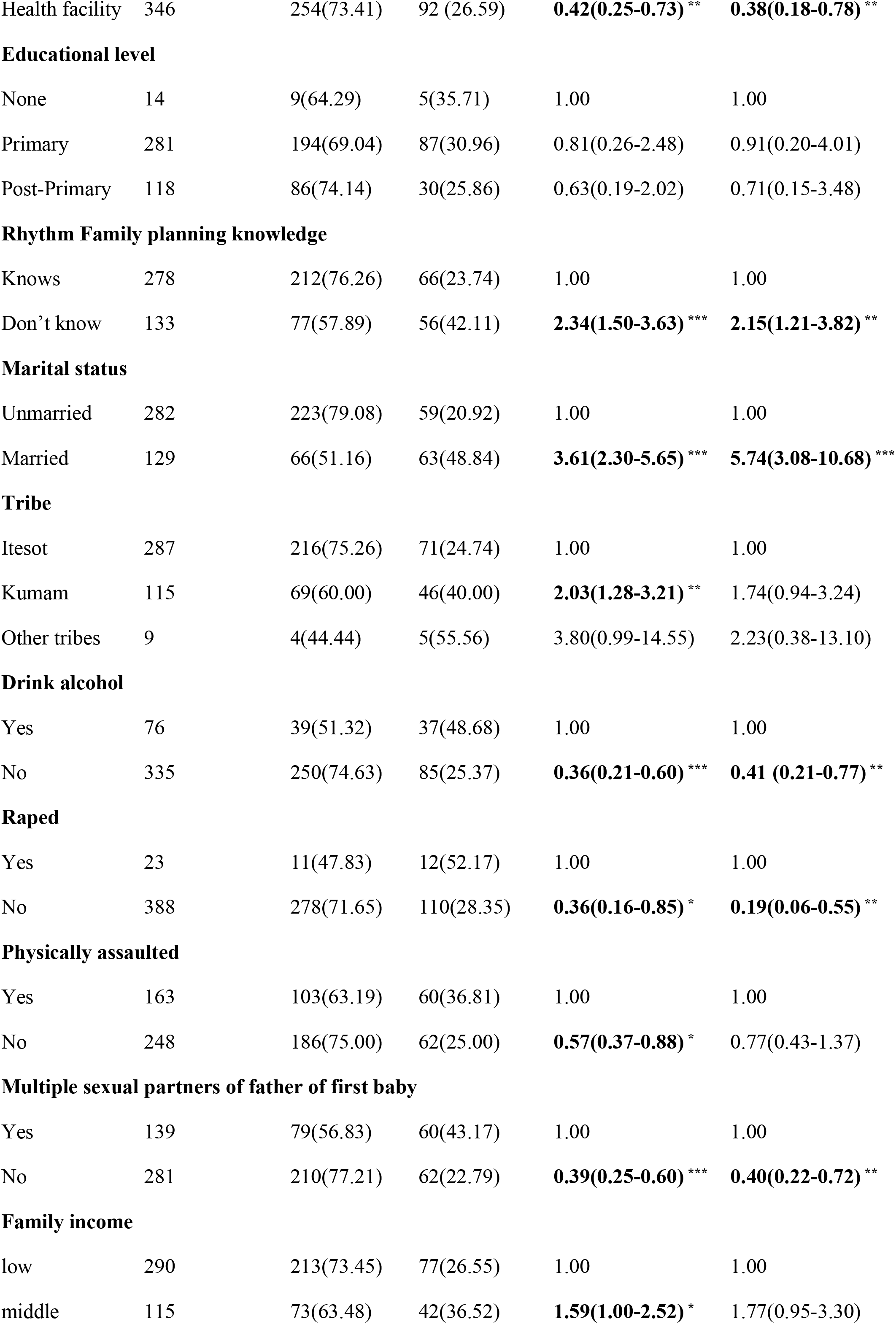
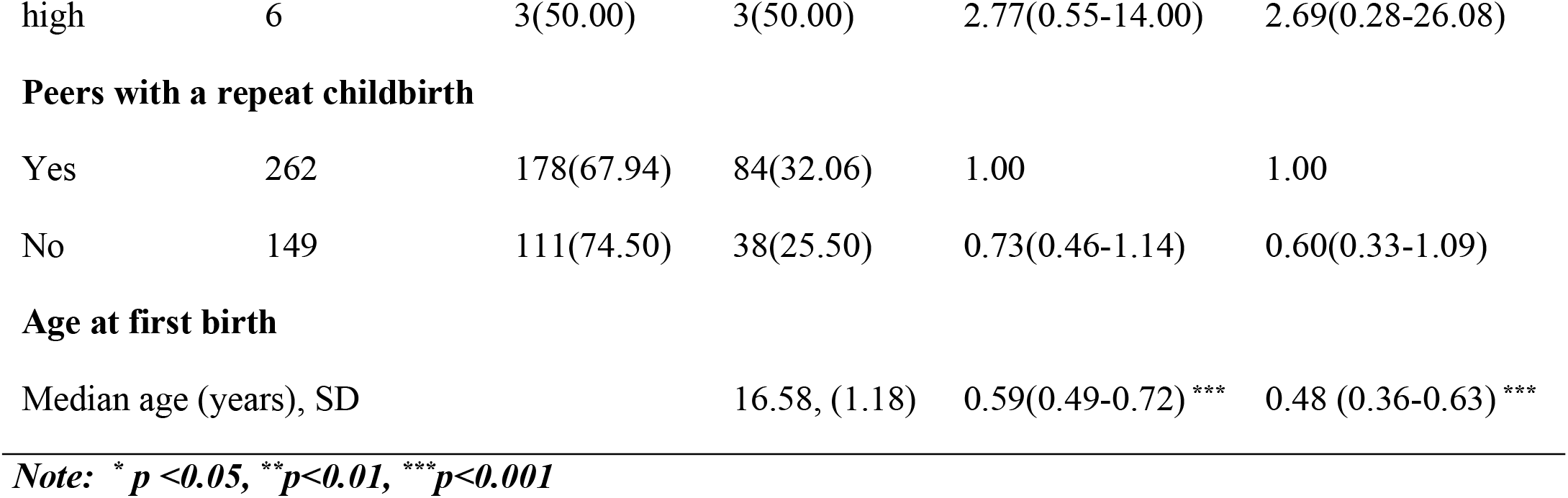
Factors associated with repeat childbirth among adolescent mothers.

## DISCUSSION OF RESULTS

We enrolled 422 respondents with the objective of determining the current proportion of mothers with, and factors associated with, repeat childbirth among adolescent mothers aged 15 to 19 in Soroti district.

### Proportion of adolescent mothers with repeat childbirth

In our study, we found a statistically high proportion of adolescent mothers with a repeat childbirth of **30.81%** in Soroti district as compared to the reported 26.1% in Uganda, or the global ARCR of 18.5% [9].

It may be difficult to compare the study findings with other similar studies done in Uganda, and in Africa, due to differences in the definition of adolescent repeat childbirth (precisely difference in the maximum age/upper limit for adolescence). For example, a study done in Uganda to estimate repeat adolescent birth defined ARC as a second or higher live birth following a first birth at <18 years among women age 20–24 years[13]. The prevalence of ARC was found to be 55.6%[13]. Another study done in Uganda among married or cohabiting women aged 15–22 with one or two previous pregnancies were drawn from the 2011 Uganda Demographic and Health Survey; rapid repeat pregnancy was defined as a pregnancy occurring within 24 months or 12 months of a prior pregnancy outcome. Of the respondents, 74% and 37% had experienced a rapid repeat pregnancy within 24 months and 12 months, respectively[37]. In the above studies, it will be noted that their participant age ranges were slightly older than ours.

To have a more meaningful comparison of repeat childbirth among adolescent mothers across countries, it is better to take a definition that consider mothers who are in adolescence. Such definitions would provide comparable findings with minimal recall bias since mothers are interviewed when they are still adolescents.

Secondly, childbirth during adolescence is associated with more poor outcomes, as compared to childbirth within other age groups. Therefore, isolating the statistics applicable to only adolescents would provide better information to guide interventions.

In a study done in 2013 in the USA to determine repeat childbirth among teens aged 15–19 years, 18.3% were repeat births[7].

A study done in South Africa in 2017 determined repeat adolescent pregnancy among females aged 13 to 19 years, as its outcome. The prevalence of repeat adolescent pregnancy was 19.9%[38]. Our study findings indicate a higher adolescent childbearing rate of 30.81% [95%CI: 26.6% - 35.4%] as compared to that in USA (18.3%) and South Africa (19.9%). The difference is statistically high upon examination of the confidence intervals.

The factors associated with repeat adolescent childbearing in Soroti district, coupled with the perceptions of adolescent mothers towards adolescent repeat childbearing are the plausible explanation for the observed high proportion of adolescent mothers with repeat childbirth in Soroti district as shown in section Factors associated with repeat childbirths among adolescent girls and

### Factors associated with repeat childbirths among adolescent girls

Married respondents were at a higher risk of adolescent repeat childbirth compared to unmarried respondents. Adolescent mothers viewed adolescent marriages as a privilege and a way of overcoming problems following their first childbirth according to the qualitative findings. Married adolescent mothers often had no say as to whether have sex or not, given that the sexual partner has demanded. In addition, childbearing among married adolescent mothers was viewed as task for which they had been taken for.

Low family income level is yet another factor that predisposes adolescent mothers in Soroti district to marriage. Qualitative results further reveal that pregnant adolescent girls were sent in for marriage in exchange for dowry, or money to support their families or pay bride price for their brothers.

Qualitative findings suggest that family and community reactions towards childbearing among adolescent mothers could yet be another reason for the observed high adolescent repeat childbearing rate in Soroti. This conforms to findings from a study done in USA in 2013 among teens aged 15 to 19 years [39, 40]. Once adolescent girls become pregnant/deliver, they often experience bad reactions from their family members inform of either physical or emotional violence or both. This increases fear among them, and they elope with their sexual partners as revealed by the qualitative findings. The likelihood of such adolescents to have a repeat childbirth is enhanced.

Other possible explanations for the observed rate of adolescent repeat childbearing may include; low sexual and reproductive health information among adolescent mothers; use of rhythm family planning method [12]), amidst inappropriate knowledge about its use leading to failure of the family planning method to work as expected as shown by the qualitative findings.

In line with a study done in Uganda, programs that increase family planning use have not been effective in Soroti district[13] amidst the well documented Adolescent Health Policy Guidelines and Services Standards that guide the National Adolescent Health Strategy.

Differences in other socio-economic and demographics of the adolescents in Soroti district and those in USA and South Africa, is yet another reason for the observed difference in adolescent childbearing rate between our findings and findings from other similar studies. For example, qualitative findings further reveal that the local council ones’(LC1s’), sexual partners and the parents of adolescent girls under 18 years conceal information from the law enforcers, once girls get pregnant. Sexual partners of the pregnant adolescent girls are often fined, as the adolescent girls are given in for marriage. Thus, the law against early marriage and defilement has many challenges regarding its enforcement.

Adolescent repeat childbirth was significantly associated with the place of delivery of the first child. Though 84.36% of the adolescent mothers delivered their first baby from a healthy facility, those who delivered at home had higher likelihood of having a repeat childbirth. These findings are similar to study done in the Philippines [41]. No related study in Uganda had reported such findings. This is because such studies did not include place of delivery in their analysis. Participants who delivered their first baby at their homes could have missed essential post-natal counselling and health education, which often is given during antenatal and postnatal visits. Such adolescent mothers could have been at high likelihood of receiving wrong information from non-health workers including the traditional birth attendants, which placed them at risk of a repeat childbirth.

Socio-cultural factors such as residence, and tribe were not significantly associated with repeat childbirth at multivariate analysis. These findings conforms to findings from a similar study done in Uganda by analyzing six demographic and health surveys[13]. Even though 93.08% of the repeat childbirths were from rural settings, residence was not significantly associated with repeat childbirth. These results differ from findings from a study done in Uganda by analyzing the 2011 demographic and health survey[40]. However, the findings are similar to another study done in Uganda by analyzing six health and demographic survey[13].

From our findings, the risk of having a repeat childbirth decreased for every one-year increase in age at first childbirth. This is in line with a study done in Uganda in 2020, where the 2016 UDHS data was analyzed [13]. Though the study population in that study differed (analyzed data for women aged 20 to 24 years), the outcome of the study (repeat adolescent childbirth) was like ours. Therefore, our study supports the findings that early childbearing among adolescent girls increases the risk of adolescent repeat childbirth. The probable explanation could be that policies and programs addressing repeat childbirth in Soroti district have not implemented measures to delay age at first childbirth among adolescents girls[19, 34]. This was evidenced by the qualitative findings where adolescent mothers confessed having received no sexual and reproductive health information before and after their first childbirth.

Our findings indicate a significant association between adolescent repeat childbirth and alcohol consumption and being raped. In the previous studies done in Uganda, no such findings were reported. This could be because alcohol consumption was not included in the analysis of such studies. However, previous studies have shown alcohol consumption to increase the risk of risky sexual behavior including; early onset of sexual activity, low condom use and having multiple sexual partners[42, 43]. In addition to the above findings from previous studies, our study reveals a significant association between alcohol consumption and religion. Compared to Catholic and Anglican respondents, respondents of other religious affiliations were at a significantly decreased risk of drinking alcohol. Urgent attention is called for, to address repeat childbirth given the high proportion of adolescent mothers at risk of having repeat childbirth (82.01% of the respondents were Catholics and Anglicans and 87.34% of the respondents who drank alcohol were also catholic and Anglicans). In addition, some of the adolescent mothers sell alcohol, which predispose them to alcohol consumption according to the qualitative findings. Perhaps, the drunkard adolescent mothers may have sexual intercourse without a clear consent since they are drunk.

Above ninety percent (91.31%) of the respondents who were raped were Catholics and Anglicans. Therefore, drinking of alcohol may increase the likelihood of being raped as evidenced from previous studies[42]. The study findings showed that adolescent mothers who were not raped were at a lower risk of a repeat childbirth. These findings complement findings from a study done in 2012 to find a place of pheromones in rape[44]. This could be attributed to the fact during ovulation in human females, pheromones are secreted by females that attract men [44]. It could also be coincidental that, at the time of rape, the adolescent mother has ovulated. Thus, rape increases the likelihood of a repeat pregnancy and its outcomes including repeat childbirth.

The partner’s characteristics associated with repeat childbirth included; the father of the first baby having multiple sexual partners. This conforms to findings from other related studies in Philippines[41], though no such findings have been reported in Uganda. This is because other related studies done in Uganda did not include sexual partner characteristics in their analysis.

## CONCLUSION AND RECOMMENDATION

The proportion of adolescent mothers aged 15 to 19 years with repeat childbirth (30.81% (95%CI: 26.57%-35.39%)) was significantly higher than the national average (26.1%) (S1 Table). This suggests urgent intervention to prevent repeat adolescent childbearing in Soroti district by the district stakeholders including the Soroti district health office.

From the results (S2 Table), the high-risk factors for repeat adolescent childbirth included having a first childbirth from home, starting childbearing at a young age, adolescent mothers who drink alcohol, adolescent marriage, and incorrect knowledge about rhythm family planning method.

This therefore suggests the following; involvement of males in preventive programs (such as family planning) of adolescent repeat childbearing since sexual partner characteristics were associated with repeat childbirth among the participants.

Also, in order to prevent repeat adolescent childbearing in Soroti district, and contribute towards the achievement of the SDG numbered three (ensure healthy lives and promote well-being for all at all ages), our study findings suggest the need to awaken and strengthen the implementation of the anti-teen marriage programs and policies; strengthen sexual/reproductive education including family planning programs, and addressing identified myths regarding ARC; and reinforce programs that ban alcohol consumption among adolescents in Soroti district by the different interventions aimed at preventing ARC and stakeholders including Soroti district health office and ministry of health. The study results further suggest the need to instate measures to delay age at first delivery among adolescent mothers so as prevent adolescent repeat childbearing by the various interventions aimed at preventing ARC in Soroti district.

We also recommend further research to be done in Uganda, especially in other districts of Teso region to validate our findings.

## Data Availability

All relevant data are within the manuscript and its Supporting Information files.

## Acknowledgement

I would like to acknowledge the various stake holders of Busitema University most especially the faculty of Health Sciences; thank you for being not only academic mentors, but also parents. Special thanks go to Prof. Peter Olupot-Olupot, Mr. Okello Francis, Dr. Matovu JKB, Dr. Iramoit Jacob, Dr. Dinah Amongin, Dr. Mukone George and the entire department of Public health, Busitema University for the scholarly guidance. In a very special way, I would like to thank Dr. Wanume Benon and Dr. Soita David for the tireless academic support they extended to this study. Gratitude goes to my wife; Joan, our children; Abigail, Ariana and Anna, for their love and support that kept me going.

## Supporting Information

Appendix

## REFERENCES

1. WHO: Adolescent pregnancy fact sheet. Adolesc. Pregnancy Fact Sheet. (2012). https://doi.org/ http://www.who.int/mediacentre/factsheets/fs364/en/

2. Charles, J.M., Rycroft-malone, J., Aslam, R., Hendry, M., Pasterfield, D.: Reducing repeat pregnancies in adolescence : applying realist principles as part of a mixed-methods systematic review to explore what works, for whom, how and under what circumstances. BMC Pregnancy Childbirth. 1–10 (2016). https://doi.org/10.1186/s12884-016-1066-x

3. Taylor, J.L.: Midlife Impacts of Adolescent Parenthood. 484–510 (2009)

4. East, P.L., Chien, N.C., Barber, J.S.: Adolescents’ Pregnancy Intentions, Wantedness, and Regret: Cross-Lagged Relations With Mental Health and Harsh Parenting Patricia. 74, 167–185 (2013). https://doi.org/10.1111/j.1741-3737.2011.00885.x.Adolescents

5. Csikszentmihalyi, M.: Adolescence, https://www.britannica.com/science/adolescence, (2020)

6. CDC: Preventing Repeat Teen Births VitalSigns CDC. (2013)

7. Gavin, L., Warner, L., O’Neil, M.E., Duong, L.M., Marshall, C., Hastings, P.A., Harrison, A.T., Barfield, W.: Vital signs: Repeat births among teens - United States, 2007-2010. Morb. Mortal. Wkly. Rep. 62, 249–255 (2013)

8. UNICEF: Early Childbearing, (2019)

9. Norton, M., Chandra-Mouli, V., Lane, C.: Interventions for preventing unintended, rapid repeat pregnancy among adolescents: A review of the evidence and lessons from high-quality evaluations, (2017)

10. WHO: Preventing Early Pregnancy and Poor Reproductive Outcomes Among Adolescents in Developing Countries. (2011)

11. McKenry, P.C., Walters, L.H., Johnson, C.: Adolescent Pregnancy: A Review of the Literature. Fam. Coord. 28, 17 (2013). https://doi.org/10.2307/583263

12. UBOS: Uganda demographic and health survey 2016. Key indicators report. (2017)

13. Amongin, D., Nakimuli, A., Hanson, C., Nakafeero, M., Kaharuza, F., Atuyambe, L., Benova, L.: Time trends in and factors associated with repeat adolescent birth in Uganda: Analysis of six demographic and health surveys. PLoS One. 15, 1–14 (2020). https://doi.org/10.1371/journal.pone.0231557

14. Azevedo, W.F. De, Diniz, M.B., Evangelista, C.B.: Complications in adolescent pregnancy : systematic review of the literature. 1–9 (2014). https://doi.org/10.1590/S1679-45082015RW3127

15. Pendse, R., Mcclure, K., Mouli, V., Health, A., Mathai, M., Portela, a: Adolescent Pregnancy. WHO MPS Notes. (2008). https://doi.org/10.1016/j.ijgo.2010.06.023

16. WHO: Pregnancy and childbirth outcomes among adolescent mothers_ a World Health Organization multicountry study - Ganchimeg - 2014 - BJOG_ An International Journal of Obstetrics &amp; Gynaecology - Wiley Online Library, (2014)

17. WHO: Adolescent pregnancy; Key facts. (2020)

18. Ganchimeg, T., Ota, E., Morisaki, N., Laopaiboon, M., Lumbiganon, P., Zhang, J., Yamdamsuren, B., Temmerman, M., Say, L., Tunçalp, Ö., Vogel, J.P., Souza, J.P., Mori, R., WHO Multicountry Survey on Maternal Newborn Health Research Network: Pregnancy and childbirth outcomes among adolescent mothers: a World Health Organization multicountry study. BJOG. (2014). https://doi.org/10.1111/1471-0528.12630

19. CDC: Preventing Repeat Teen Births

20. Gonçalves, S.D., Moultrie, T.A.: Short preceding birth intervals and child mortality in Mozambique. Afr. J. Reprod. Health. 16, 29–42 (2012)

21. Grundy, E., Kravdal, Ø.: Do short birth intervals have long-term implications for parental health? Results from analyses of complete cohort Norwegian register data. J. Epidemiol. Community Health. 68, 958–964 (2014). https://doi.org/10.1136/jech-2014-204191

22. de Jonge, H.C.C., Azad, K., Seward, N., Kuddus, A., Shaha, S., Beard, J., Costello, A., Houweling, T.A.J., Fottrell, E.: Determinants and consequences of short birth interval in rural Bangladesh: A cross-sectional study. BMC Pregnancy Childbirth. 14, 1–7 (2014). https://doi.org/10.1186/s12884-014-0427-6

23. Thompson, D.R., Clark, C.L.: Short Birth Intervals : Associated Maternal Factors and Subsequent Risk of Adverse Birth Outcomes Division of Family Health Services. 1–12 (2011)

24. Govender, D., Naidoo, S., Taylor, M.: Scoping review of risk factors of and interventions for adolescent repeat pregnancies: A public health perspective. African J. Prim. Heal. Care Fam. Med. 10, (2018). https://doi.org/10.4102/phcfm.v10i1.1685

25. Yogman, M., Garfield, C.F.: Fathers’ roles in the care and development of their children: The role of pediatricians. Pediatrics. 138, (2016). https://doi.org/10.1542/peds.2016-1128

26. De Freitas Galvão, R.B., Figueira, C.O., Borovac-Pinheiro, A., De Morais Paulino, D.S., Faria-Schützer, D.B., Surita, F.G.: Hazards of repeat pregnancy during adolescence: A case-control study. Rev. Bras. Ginecol. e Obstet. 40, 437–443 (2018). https://doi.org/10.1055/s-0038-1666811

27. Uganda Bureau of Statistics: National Population and Housing Census 2014 - Main Report. Uganda Bur. Stat. (2014). https://doi.org/10.1017/CBO9781107415324.004

28. Uganda Bureau of Statistics: National Population and Housing Census 2014 - Main Report. Uganda Bur. Stat. 1–209 (2014). https://doi.org/10.1017/CBO9781107415324.004

29. The Alan Guttmacher Institute: Research In Brief Adolescents in Uganda : (2001)

30. Poedjastoeti, S., Cross, A.: Reproductive Health of Young Adults in Uganda. 60 (2001)

31. Slaymaker, E., Bwanika, J.B., Kasamba, I., Lutalo, T., Maher, D., Todd, J.: Trends in age at first sex in Uganda: Evidence from Demographic and Health Survey data and longitudinal cohorts in Masaka and Rakai. Sex. Transm. Infect. 85, (2009). https://doi.org/10.1136/sti.2008.034009

32. Jones, P.W., Coleman, D.: UNDP. In: The United Nations and Education. pp. 187–227 (2010)

33. Meckstroth, A., Berger, A.: Preventing Rapid Repeat Teen Pregnancy with Motivational Interviewing and Contraceptive Access : Implementing Teen Options to Prevent Pregnancy (T. O. P. P.) Implementation Report This page is left blank for double-sided printing. (2014)

34. Ministry of Health: Adolescent Health Policy Guidelines and Service Standards. (2012)

35. Amin, S., Karen Austrian, Michelle Chau, Kimberly Glazer, Eric Green David Stewart, M.S.: THE ADOLESCENT GIRLS VULNERABILITY INDEX: GUIDING STRATEGIC INVESTMENT IN UGANDA, (2013)

36. Population, N., Profiles, A.S.: Area Specific Profiles Soroti District. (2017)

37. Mcclain, H., Santo, L.D., Bernholc, A., Akol, A., Dal, L., Burke, H.M.: Correlates of Rapid Repeat Pregnancy Among Adolescents and Young Women in Uganda. 44, 11–18 (2018)

38. Govender, D., Naidoo, S., Taylor, M.: Prevalence and Risk Factors of Repeat Pregnancy among South African Adolescent Females. Afr. J. Reprod. Health. (2019). https://doi.org/10.29063/ajrh2019/v23i1.8

39. Boardman, L.A., Sc, M., Allsworth, J., Ph, D., Phipps, M.G., h, M.P., Lapane, K.L., Ph, D.: Risk Factors for Unintended Versus Intended Rapid Repeat Pregnancies among Adolescents. 39, 1–8 (2006). https://doi.org/10.1016/j.jadohealth.2006.03.017

40. Gideon, R.: Factors Associated with Adolescent Pregnancy and Fertility in Uganda: Analysis of the 2011 Demographic and Health Survey Data. Soc. Sci. (2013). https://doi.org/10.5923/j.sociology.20130302.03

41. Maravilla, J.C., Betts, K.S., Alati, R.: Exploring the Risks of Repeated Pregnancy Among Adolescents and Young Women in the Philippines. Matern. Child Health J. 23, 934–942 (2019). https://doi.org/10.1007/s10995-018-02721-0

42. Bellis, M.A., Morleo, M., Tocque, K., Dedman, D., Clare, P.P., Lisa, P.: CONTRIBUTIONS OF ALCOHOL An initial examination of geographical and evidence based associations.

43. Youth, M., Behavior, R.: Alcohol Consumption and Teen Pregnancy 2013. 42, 2012–2013 (2013)

44. Skandhan, K.P., Kaoru, O., Mukund, B.M., Sumangala, B.: Place of Pheromone in Rape. 2013, 24–27 (2013). https://doi.org/10.4236/asm.2013.31005

